# Molecular pathology and synaptic loss in primary tauopathies: [^18^F]AV-1451 and [^11^C]UCB-J PET study

**DOI:** 10.1101/2021.03.03.21252808

**Authors:** Negin Holland, Maura Malpetti, Timothy Rittman, Elijah E Mak, Luca Passamonti, Sanne S Kaalund, Frank H. Hezemans, P. Simon Jones, George Savulich, Young T. Hong, Tim D. Fryer, Franklin I. Aigbirhio, John T O’Brien, James B Rowe

## Abstract

The relationship between *in vivo* synaptic density and tau burden in primary tauopathies is key to understanding the impact of tauopathy on functional decline and in informing new early therapeutic strategies. In this cross-sectional observational study, we determine the *in vivo* relationship between synaptic density and molecular pathology, in the primary tauopathies of Progressive Supranuclear Palsy (PSP) and Corticobasal Degeneration (CBD), as a function of disease severity.

Twenty three patients with PSP, and twelve patients with Corticobasal Syndrome (CBS) were recruited from a tertiary referral centre. Nineteen education, sex and gender-matched control participants were recruited from the National Institute for Health Research ‘Join Dementia Research’ platform. Cerebral synaptic density and molecular pathology, in all participants, were estimated using PET imaging with the radioligands [^11^C]UCB-J and [^18^F]AV-1451, respectively. Patients with CBS also underwent amyloid PET imaging with [^11^C]PiB to exclude those with likely Alzheimer’s pathology – we refer to the amyloid negative cohort as having CBD although acknowledge other pathologies exist. Disease severity was assessed with the PSP rating scale; regional non-displaceable binding potentials (BP_ND_) of [^11^C]UCB-J and [^18^F]AV-1451 were estimated in regions of interest from the Hammersmith Atlas, excluding those with known off-target binding for [^18^F]AV-1451. As an exploratory analysis, we also investigated the relationship between molecular pathology in cortical brain regions, and synaptic density in connected subcortical areas.

Across brain regions, there was a *positive* correlation between [^11^C]UCB-J and [^18^F]AV-1451 BP_ND_ *(β*=*0*.*4, t=4*.*7, p*<*0*.*0001*). However, the direction of this correlation became less positive as a function of disease severity in patients *(β = -0*.*03, T = -4*.*0, p = 0*.*002*). Between brain regions, cortical [^18^F]AV-1451 binding was *negatively* correlated with synaptic density in subcortical areas (caudate nucleus, putamen, and substantia nigra).

Brain regions with higher synaptic density are associated with a higher [^18^F]AV-1451 binding in PSP/CBD, but this association diminishes with disease severity. Moreover, higher cortical [^18^F]AV-1451 binding correlates with lower subcortical synaptic density. Longitudinal imaging is required to confirm the mediation of synaptic loss by molecular pathology. However, the effect of disease severity suggests a biphasic relationship between synaptic density and tauopathy, with synapse rich regions vulnerable to accrual of pathology, followed by a loss of synapses in response to pathology. Given the importance of synaptic function for cognition, our study elucidates the pathophysiology of primary tauopathies and may inform the design of future clinical trials.

## Introduction

Synaptic loss is a feature of many neurodegenerative disorders^1–3^. It is closely related to cognitive decline in symptomatic stages of disease^4,5^, but can begin long before symptom onset and neuronal loss^6^. Synaptic loss and dysfunction may be an important mediator of decline even where atrophy is minimal or absent^7,8^. Conversely, synaptic connectivity may facilitate the spread of oligomeric mis-folded proteins such as tau^9,10^. The relationship between synaptic loss and the accumulation of mis-folded proteins in primary tauopathies has yet to be determined *in vivo*. Preclinical models suggest early synaptotoxicity of oligomeric tau, leading to reduced plasticity and density^11,12^. In patients with mutations of microtubule-associated protein tau (MAPT), there are deficiencies in many synaptic pathways including GABA-mediated signalling and synaptic plasticity^13^. The mechanisms of synapse loss following tau pathology include both direct and indirect pathways (reviewed in Spires-Jones *et al. 2014*^14^). In the related tauopathy of Alzheimer’s disease, there is differential expression of synaptic proteins in the early stages^15,16^; this may be an attempt to maintain cellular physiology, which fails as the disease progresses, leading to loss of synaptic function and synapse numbers.

In clinical disorders, the *in vivo* pathologies of synaptic density and tau burden can be characterised by positron emission tomography (PET). In Alzheimer’s disease for example, increased temporal lobe binding of the tau radioligand [^18^F]MK-6240 is associated with decreased synaptic density measured by the radioligand [^11^C]UCB-J^17^. However, the pathology of Alzheimer’s disease is multifaceted with amyloid and tau aggregation, vascular changes and neuroinflammation^18^.

In this study, we use Progressive Supranuclear Palsy (PSP) and Corticobasal Degeneration (CBD) as models of human tauopathy, with relevance to other tau-mediated neurodegenerative disorders, and examine the relationship between synaptic density and tau burden^19^. An advantage of studying PSP is the very high correlation between the clinical syndrome, and the specific 4R-tauopathy at autopsy^20^. The clinical phenotype of Corticobasal Syndrome (CBS), may be caused by CBD, but can also be mimicked by Alzheimer’s disease, and less commonly by other forms of frontotemporal lobar degeneration^20^. Here, we use the term CBD to refer to patients with CBS in whom Alzheimer’s disease is excluded by [^11^C]PiB PET, whereby in the absence of amyloid pathology there is a high clinicopathological correlation with 4R-tauopathy at post mortem. Both PSP and CBD demonstrate synaptic loss *in vivo*^7,8^ and at post-mortem^1,2^. The distribution of tau pathology in both diseases is well characterised with cortical and subcortical involvement^21,22^. Animal models of tauopathy have illustrated the colocalisation of misfolded tau protein and synaptic loss at the synaptic bouton^14,23^ but the tau-synapse association is yet to be determined *in vivo*.

Figure 1 illustrates our hypotheses. Previous studies suggest that the strength of connectivity within a region and between brain regions can promote the spread of tau pathology, in humans as in preclinical models^10,24^. Therefore, we hypothesised that brain areas with higher synaptic density would develop more tau pathology. We predicted that the spatial distribution of pathology, as measured with the PET radioligand [^18^F]AV-1451, would be correlated with synaptic density, as measured with the PET radioligand [^11^C]UCB-J (which binds to the presynaptic vesicle glycoprotein SV2A that is ubiquitously expressed in all brain synapses^25,26^). Since tauopathy in a region may impair efferent projections, a corollary hypothesis is that tau accumulation in one region (source region) leads to diaschisis characterised by reduced synaptic density in the areas to which it connects (target regions).

**Figure 1.**
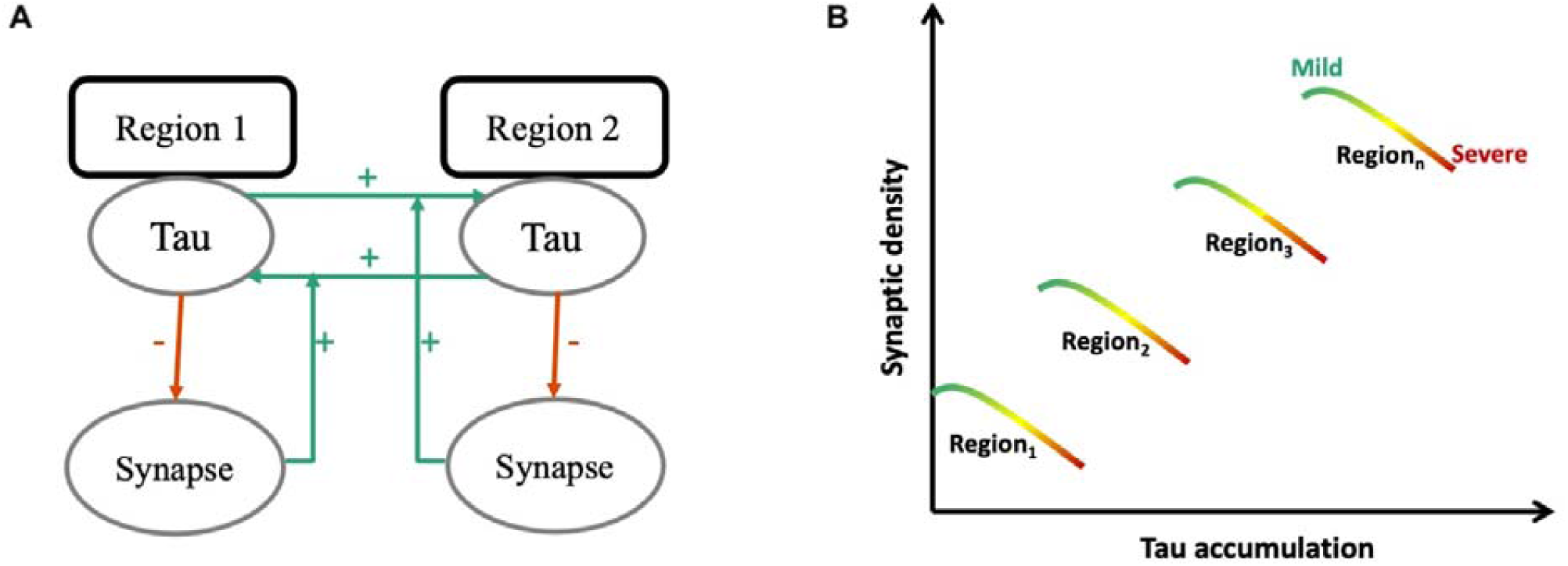
Schematic diagram illustrating the predicted toxic effect of tau on synaptic density. At a regional level (A) synaptic density promotes the spread of tau from one region to another. Tau burden therefore depends on a region’s baseline synaptic density, for example in B, region 3 would accumulate more tau given its high baseline synaptic density. However, this relationship between tau and synapses is further affected by stage of disease (B) such that in any given region, tau-induced synaptic loss ensues as the stage of disease progresses from mild to moderate to severe.

We acknowledge the relatively low affinity of [^18^F]AV-1451 for 4R tauopathy compared to Alzheimer’s disease, and the off-target binding of this ligand within the basal ganglia. We therefore refer to its binding target as ‘molecular pathology’, covering tau and non-tau targets.

A second part of the model describes the consequence of the pathology, which is to reduce synaptic density. The predicted result is a positive relationship between [^18^F]AV-1451 binding and synaptic loss, negatively moderated by disease progression (Figure 1b).

## Materials and Methods

### Participant recruitment and study design

Twenty three people with probable PSP–Richardson Syndrome^19^, and twelve people with probable CBS in whom Alzheimer’s disease was excluded with [^11^C]PiB PET ^7^, were recruited from a regional specialist National Health Service clinic at the Cambridge University Centre for Parkinson-plus. We refer to our amyloid-negative CBS cohort as having CBD but acknowledge other pathologies are possible^20^. Nineteen healthy volunteers were recruited from the UK National Institute for Health Research Join Dementia Research (JDR) register. Participants were screened using the inclusion/exclusion criteria set out in Holland *et al. 2020*^7^. Eligible participants underwent clinical and cognitive assessments (Table 1) including the revised Addenbrooke’s Cognitive Examination (ACE-R), and the mini-mental state examination (MMSE); disease severity was measured with the PSP rating scale. Participants underwent 3T MRI, [^18^F]AV-1451 PET, and [^11^C]UCB-J PET. The research protocol was approved by the Cambridge Research Ethics Committee (reference 18/EE/0059) and the Administration of Radioactive Substances Advisory Committee. All participants provided written informed consent in accordance with the Declaration of Helsinki.

**Table 1.**
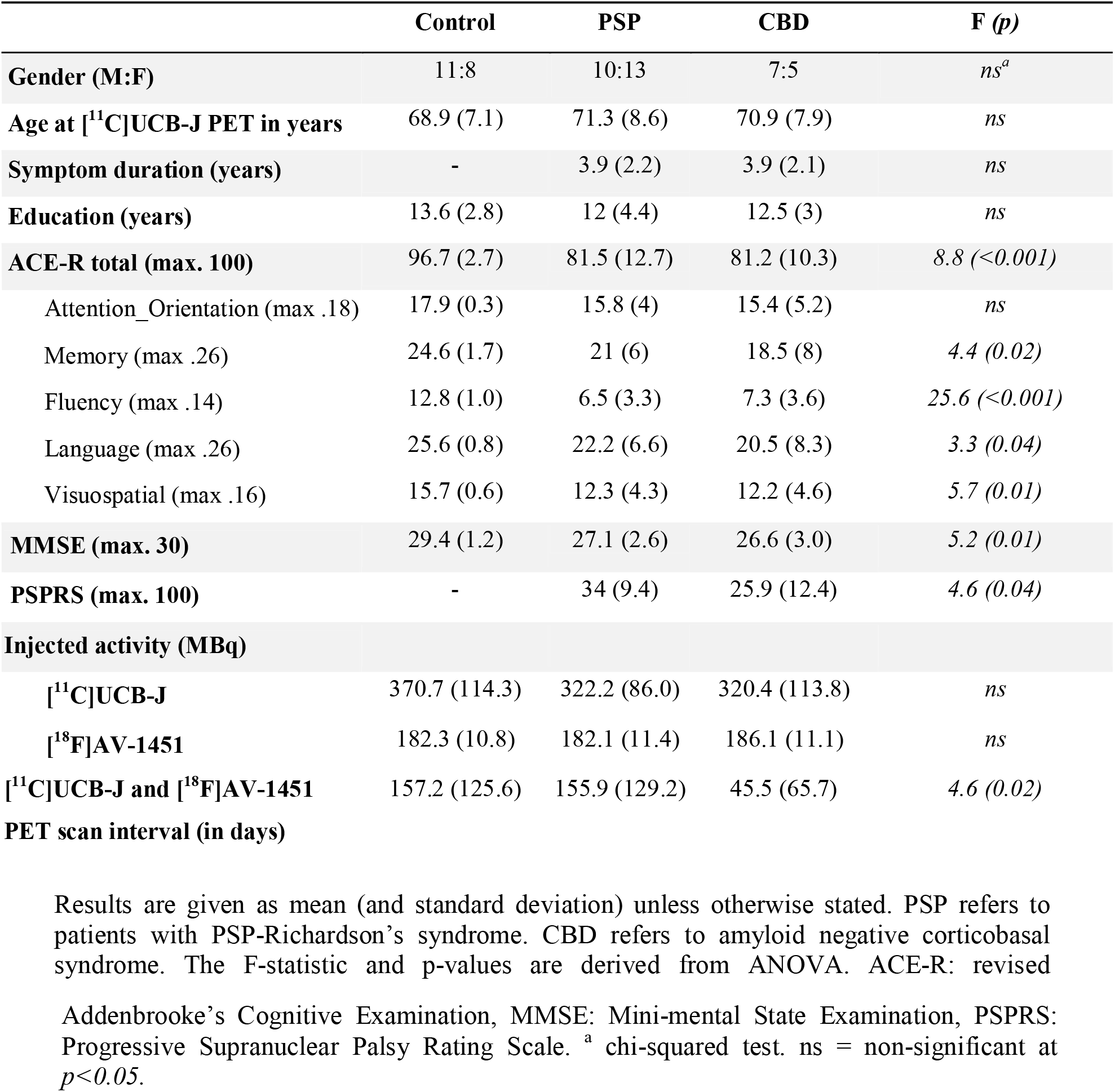
Clinical and Demographics summary.

### PET data acquisition and kinetic analysis

#### [^11^C]UCB-J PET

The procedure for [^11^C]UCB-J synthesis, PET data acquisition, image reconstruction and kinetic analysis was the same as in Holland *et al*. 2020^7^. In brief, dynamic PET data acquisition was performed on a GE SIGNA PET/MR (GE Healthcare, Waukesha, USA) for 90 minutes immediately after injection, with attenuation correction using a multi-subject atlas method ^27,28^ and improvements to the MRI brain coil component^29^. Emission image series were aligned using SPM12 (www.fil.ion.ucl.ac.uk/spm/software/spm12/), and rigidly registered to the T1-weighted MRI acquired during PET data acquisition (TR = 3.6 msec, TE= 9.2 msec, 192 sagittal slices, in plane resolution 0.55 × 0.55 mm, interpolated to 1.0 × 1.0 mm; slice thickness 1.0 mm). The Hammersmith atlas (http://brain-development.org) with modified posterior fossa regions was spatially normalized to the T1-weighted MRI of each participant using Advanced Normalisation Tools (ANTs) software^30^. Regional time-activity curves were extracted following the application of geometric transfer matrix (GTM) partial volume correction (PVC ^31^) to each dynamic PET image. Regions of interest (ROIs) were multiplied by a binary grey matter mask (>50% on the SPM12 grey matter probability map smoothed to PET spatial resolution), with the exception of the subcortical grey matter regions pallidum, substantia nigra, pons and medulla. To assess the impact of PVC, time-activity curves were also extracted from the same ROIs without the application of GTM PVC (discussed below as “without partial volume correction”).

To quantify SV2A density, [^11^C]UCB-J non-displaceable binding potential (BP_ND_) was determined using a basis function implementation of the simplified reference tissue model^32^, with the reference tissue defined in the centrum semiovale^33,34^.

#### [^18^F]AV-1451 PET

[^18^F]AV-1451 synthesis and data acquisition followed the protocol given in Passamonti et al. 2018^35^, except that the data were acquired on a GE SIGNA PET/MR. [^18^F]AV-1451 BP_ND_ used the inferior cerebellum as the reference region.

#### [^11^C]PiB PET

Amyloid imaging using Pittsburgh Compound B ([^11^C]PiB) followed the protocol given in Holland et al 2020^7^. [^11^C]PiB cortical standardised uptake value ratio (SUVR; 50-70 minutes post injection) was calculated using the whole cerebellum reference tissue as per the Centiloid Project methodology^36^. A negative amyloid status was characterised by a cortical [^11^C]PiB SUVR less than 1.21 (obtained by converting the Centiloid cut-off of 19 to SUVR using the Centiloid-to-SUVR transformation)^37^.

### Statistical analyses

We compared demographic and clinical variables between the diagnostic groups using ANCOVA, and chi-square tests where appropriate. We used a linear mixed effects model to assess the overall relationship between [^18^F]AV-1451 and [^11^C]UCB-J BP_ND_, and the effect of group (patients vs controls) and brain region on this relationship. For this analysis regions of interest with previously reported off-target binding of [^18^F]AV-1451 (basal ganglia, and substantia nigra ^38^) were excluded. To investigate the effect of individual variability on the relationship between [^11^C]UCB-J and [^18^F]AV-1451 BP_ND_, we used a linear mixed effects model, allowing for an uncorrelated random slope and intercept per individual. We subsequently extracted the slope of [^11^C]UCB-J on [^18^F]AV-1451 for each individual and used this in a linear model with the PSP rating scale (a measure of disease severity) as the independent variable, and age as a covariate of no interest. To explore the correlation between [^11^C]UCB-J and [^18^F]AV-1451 BP_ND_ between regions, we calculated a correlation matrix between cortical [^18^F]AV-1451 binding and synaptic density in cortical and subcortical regions.

Analyses were performed with and without partial volume correction, yielding similar results; we focus on partial volume corrected BP_ND_ to limit the potential effect of atrophy on our ligand cross-correlation but present data without partial volume correction in the supplementary material (Supplementary Figure 1 and 2). Statistical analyses were implemented in R (version 3.6.2).

### Data Availability Statement

The data that support the findings of this study are available from the corresponding author, upon reasonable request for academic (non-commercial) purposes, subject to restrictions required to preserve participant confidentiality.

## Results

### Demographics

The patients (PSP and CBD) and control groups were similar in age, sex, education and injected activity of [^11^C]UCB-J and [^18^F]AV-1451 (Table 1). We observed typical cognitive profiles for people with PSP and CBD: impaired on verbal fluency, memory and visuospatial domains of the ACE-R and MMSE.

### Relationship between [^11^C]UCB-J BP_ND_ and [^18^F]AV-1451 BP_ND_

There was a positive relationship between [^18^F]AV-1451 BP_ND_ and [^11^C]UCB-J BP_ND_ across all participants (i.e. PSP, CBD and controls included) *(β*=*0*.*4, t=4*.*7, p*<*0*.*0001*). There were no interaction effect of group-by-[^18^F]AV-1451, or group-by-region-by-[^18^F]AV-1451. However, there was a significant region-by-[^18^F]AV-1451 interaction (*p= 0*.*002*) driven by subregions of the frontal, parietal, temporal and occipital lobes, as well as the thalamus and brainstem. The positive relationship between [^18^F]AV-1451 BP_ND_ and [^11^C]UCB-J BP_ND_ remained when running the analysis in controls *(β*=*0*.*6, t=4, p*<*0*.*0001*) and patients *(β*=*0*.*4, t=4, p= 0*.*0002*), separately. In patients alone there was no interaction effect of patient group (PSP/CBD)-by-[^18^F]AV-1451 or group-by-region-by-[^18^F]AV-1451, but a significant effect of region-by-[^18^F]AV-1451 as described above.

Across all patients and regions there was a significant positive relationship between [^18^F]AV- 1451 BP_ND_ and [^11^C]UCB-J BP_ND_ *(β*=*1, T= 7, p*<*0*.*0001*). There was individual variability in the slope of this relationship (individual grey lines in Figure 2A).

The direction of the relationship between [^18^F]AV-1451 BP_ND_ and [^11^C]UCB-J BP_ND_ within each individual (i.e. the slope of each grey line in Figure 2A) negatively correlated with disease severity *(β =-0*.*03, T=-4*.*0, R=-0*.*53, p= 0*.*002*), independent of age (effect of age: *β*=*0*.*03, T= 3, p= 0*.*002*) (Figure 2B). In other words, those patients with more severe disease displayed a less positive relationship between [^18^F]AV-1451 BP_ND_ and [^11^C]UCB-J BP_ND_. Practically identical findings were observed using BP_ND_ derived from data without partial volume correction (Supplementary Figure 1). Of note, the significance of the overall model above did not change with the addition of scanning interval as a covariate of no interest.

**Figure 2.**
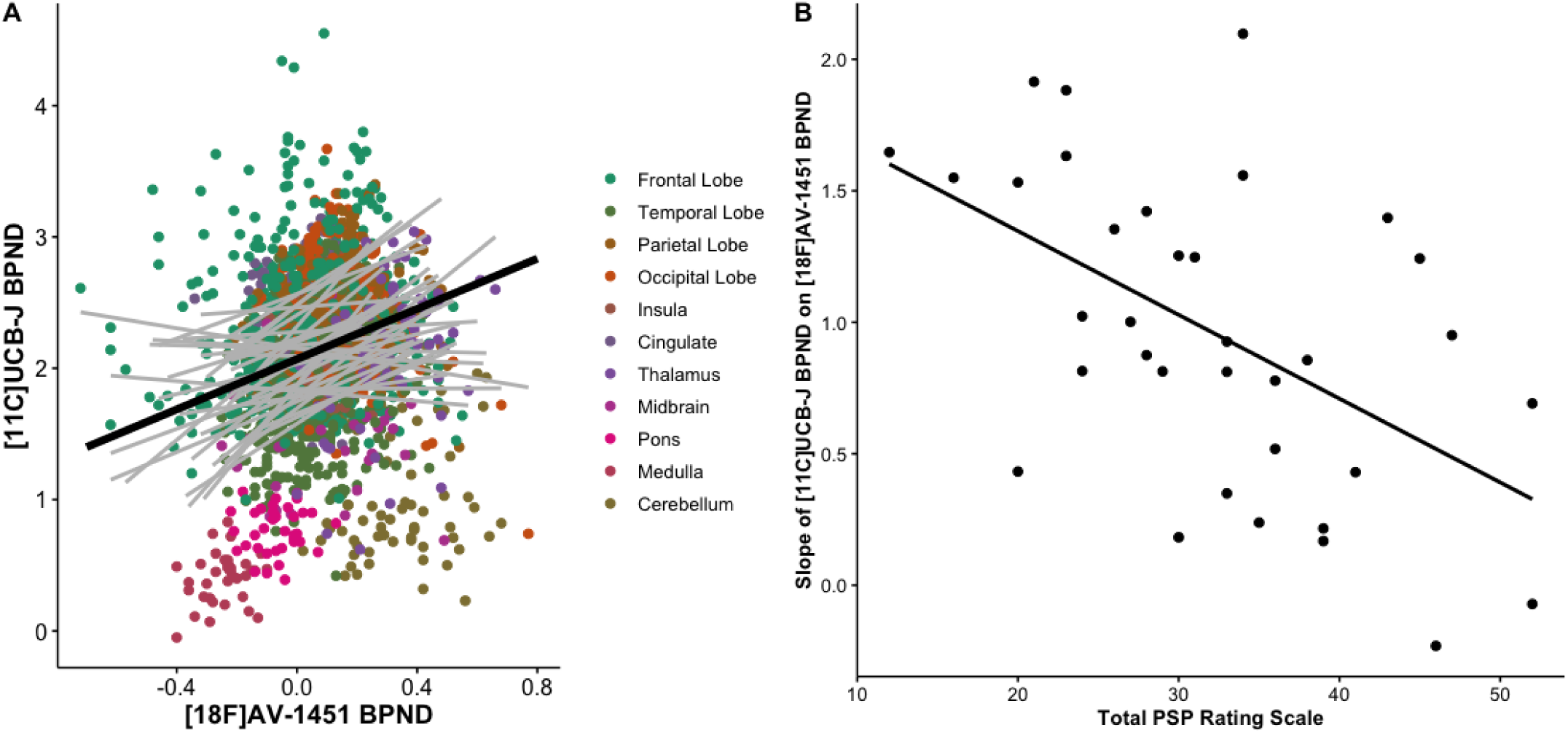
The association between synaptic density ([^11^C]UCB-J) and molecular pathology ([^18^F]AV- 1451) is a function of disease severity. A) Scatter plot of [^11^C]UCB-J BP_ND_ and [^18^F]AV-1451 BP_ND_ from 35 patients with PSP-Richardson’s syndrome and amyloid-negative CBD (each grey line represents a patient), across 73 regions of interest (excluding those with previously reported off-target binding, i.e. basal ganglia and substantia nigra); the dark black line in A depicts the overall fit of the linear mixed model, whilst grey lines represent individual patient participants. B) The slope for each individual (i.e. each grey line in A) is negatively correlated with disease severity (as measured with the PSP rating scale); *R= -0*.*53, p*<*0*.*002*.

### Cross-regional correlation between [^18^F]AV-1451 BP_ND_ and [^11^C]UCB-J BP_ND_

Synaptic density in a region is proposed to be affected by both local tau pathology and tau burden in connected regions from which it receives afferent projections. As a result, despite a positive correlation at a regional level, the synaptic density in any given region may be negatively affected by remote insult, with diaschisis between anatomically connected regions (illustrated schematically in Figure 1A). As an exploratory analysis, we computed the asymmetric Pearson’s correlation matrix shown in Figure 3, between cortical [^18^F]AV-1451 BP_ND_ (horizontal axis of matrix) and [^11^C]UCB-J BP_ND_ (vertical axis of matrix) in cortical and subcortical regions in patients. We show that overall, there are significant negative correlations between cortical (frontal, temporal, and parietal) [^18^F]AV-1451 BP_ND_ and subcortical [^11^C]UCB-J BP_ND_ within the basal ganglia and brainstem. We observe a strong positive correlation between [^18^F]AV-1451 BP_ND_ and [^11^C]UCB-J BP_ND_ within the thalamus where strong local connections exist (Figure 3).

**Figure 3.**
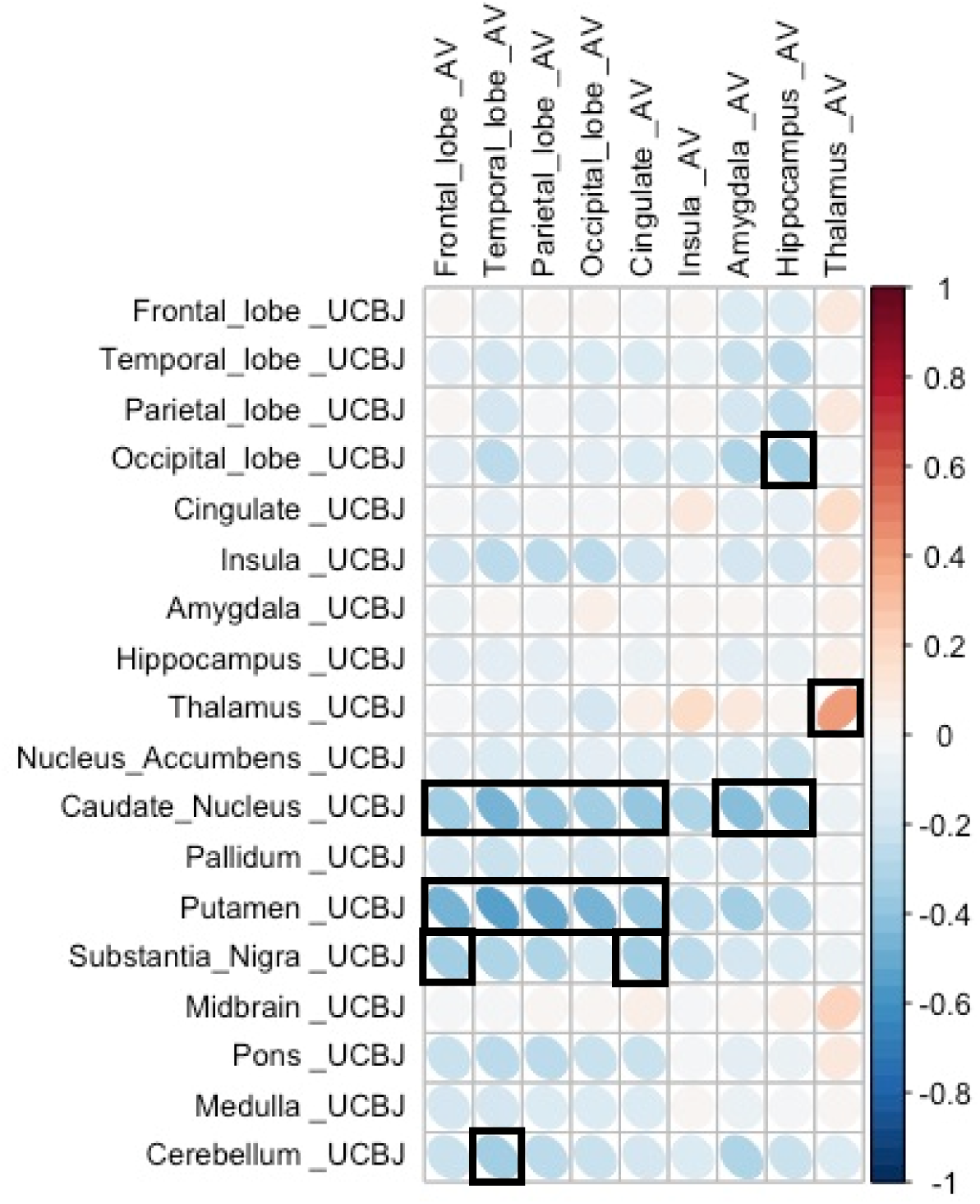
Cortical pathology is negatively correlated with subcortical synaptic density. Correlation between [^18^F]AV-1451 BP_ND_ in cortical regions (horizontal axis) and [^11^C]UCB-J BP_ND_ in a target region (vertical axis) both cortically and subcortically **in patients**. Significant correlations (at p<0.05 uncorrected for multiple comparisons) are outlined in black.

We did not include subcortical [^18^F]AV-1451 BP_ND_ in the matrix in Figure 3 given the off- target binding in these regions which undermines the interpretability of the signal. However, we include these regions as well as other subregions in the larger correlation matrix in Supplementary Figure 3 for completeness. Similar findings are seen using BP_ND_ from data without partial volume correction (Supplementary Figure 2).

## Discussion

We have identified the relationship between molecular pathology (estimated with [^18^F]AV- 1451 PET) and synaptic density (estimated with [^11^C]UCB-J PET), in patients with the primary tauopathies of Progressive Supranuclear Palsy and Corticobasal Degeneration (inferred *in vivo* from amyloid-negative corticobasal syndrome). There are three principal results: (i) regions with higher synaptic density have higher pathology, (ii) within regions, synaptic density becomes less dependent on [^18^F]AV-1451 binding as disease severity increases, and (iii) between regions, increased cortical [^18^F]AV-1451 binding is associated with reduced subcortical synaptic density. We interpret these three findings in the context of connectivity-based susceptibility to tauopathy, synaptotoxic effects of tauopathy, and cortico-subcortical diaschisis, respectively.

The effect of 4R hyperphosphorylated tau pathology such as PSP and CBD ^22,39^, on synaptic function and density is complex. It involves both direct and indirect pathways of injury with changes in cellular physiology preceding the loss of neurons. Through direct pathways, pathological tau interferes with dendritic morphology, synaptic protein expression, the number of NMDA (N-methyl-D-Aspartate) and AMPA (α-amino-3-hydroxy-5-methyl-4- isoxazolepropionic acid) receptors on the pre-synaptic membrane, mitochondrial function, synaptic vesicle numbers, and ultimately synaptic loss (for a review of animal studies illustrating various direct tau-induced synaptic abnormalities see ^40^). Tau also directly affects the axon cytoskeleton and trafficking, as well as the soma ^41^. Indirectly, hyperphosphorylated tau adversely affects the functioning of the neuronal support network, including glia cells and astrocytes ^42,43^. These events are affected by the stage and severity of the disease process, and in relation to regional differences in connectivity which we discuss next (concepts schematically illustrated in Figure 1).

We identified a positive relationship between the binding of [^11^C]UCB-J and [^18^F]AV-1451 such that areas of the brain with higher synaptic density develop higher pathology. This accords with preclinical and clinical models of tauopathy in which the strength of local network connectivity facilitates the transneuronal spread of tau pathology ^10,24,44,45^.

However, the relationship between tau accumulation and synaptic density changes with disease progression, at least as inferred from the cross-sectional moderation by disease severity (Figure 2B). With increasing scores on the PSP rating scale, synaptic density becomes less dependent on local tau accumulation. In other words, in areas with relatively low tau accumulation synaptic density is minimally affected, whereas in areas with higher tau accumulation there is reduction of synaptic density as the disease progresses and this preferentially occurs in synapse rich areas. As the disease progresses, other pathological processes may contribute to synaptic loss, such as inflammation, another predictor of prognosis and mediator of synaptic loss ^46^. There is therefore not a simple linear relationship between tau accumulation and synaptic density in moderate and advanced disease. This observation accords with human post-mortem and animal studies. In post mortem studies of the tauopathy Alzheimer’s disease, there is a biphasic synaptic protein response during disease progression, with increases in synaptophysin/syntaxin/SNAP-25 in early Braak stages and synaptic loss observed only when the disease has progressed to the neocortex ^16^. In the P301L transgenic mouse model of PSP-like tauopathy, there is a differential loss of synapses, as well as synaptic proteins, depending on disease stage^15^.

To understand the biphasic relationship between tau accumulation and synaptic density, one must consider other key players in synaptotoxicity in tauopathies, such as neuroinflammation ^47^. Recent *in vivo* studies have confirmed the regional co-localisation of inflammation and [^18^F]AV-1451 binding in PSP ^48^, in line with previous *in vivo* ^49,50^, and post mortem^51^ reports of the tight interplay between neuroinflammation and tau accumulation in tauopathies. There is growing evidence that these two pathological processes affect synaptic function both independently and synergistically ^40,42^.

The relationship between tauopathy and synaptic density is even more intriguing when considering the change in synaptic density in one region as a function of pathology in another. There are strong correlations between [^11^C]UCB-J binding within the basal ganglia and brain stem and [^18^F]AV-1451 binding in most cortical areas, with the association between synaptic density in the caudate, putamen, and substantia nigra, and cortical tau remaining significant at p<0.05. The reverse association, between subcortical [^18^F]AV-1451 and cortical [^11^C]UCB-J binding is also observed (Supplementary Figure 3) but is dismissed here as uninterpretable in view of subcortical off-target binding of [^18^F]AV-1451. The significant negative correlation between cortical [^18^F]AV-1451 binding and synaptic density in the basal ganglia could be a reflection of severe disease in the basal ganglia and accumulating pathology in the neocortex. In other words, synapses are severely affected in the basal ganglia as one of the earliest sites of pathology, with pathology spreading and accumulating in synapse-rich areas of the brain, for example the neocortex. A second explanation is that loss of descending cortico-striatal axons due to cortical pathology, may cause diaschisis, affecting subcortical synaptic density even further. Previous analysis of diffusion tensor imaging in patients with PSP/CBD have revealed extensive white matter abnormalities (within the main association fibres) beyond the degree of cortical atrophy ^52,53^ resulting in loss of cortical afferents onto subcortical structures. A third, though not mutually exclusive, explanation is the weakening of cortical-subcortical functional connectivity resulting from dysfunctional synapses rather than synaptic loss ^24^.

Although at a regional level there is a positive correlation between [^11^C]UCB-J and [^18^F]AV- 1451 BP_ND_, we are not directly measuring either synaptic function or the synaptotoxic tau oligomers. This caveat must be borne in mind when interpreting PET data. It is the preclinical models that have shown that oligomers of tau are toxic to synaptic function, even in the absence of tau polymers/fibrils ^11,12^. By the time tau aggregates are established, oligomers of tau are expected cortically, and perhaps interfering with cortical function and the integrity of descending axons.

There are other limitations to our study. First, the low affinity of [^18^F]AV-1451 for PSP and CBD 4R tau. Even where this radioligand recapitulates the distribution of post-mortem neuropathology in PSP and CBD, and binds PSP 4R tau, the affinity is very much lower than for 3R tau in Alzheimer’s disease ^21,39^. Second, there is well-established off-target binding of [^18^F]AV-1451, particularly within subcortical structures where monoamine oxidase is present. Off-target binding is most prominent in the basal ganglia which we excluded before running our statistical analyses. We included these regions in the detailed descriptive correlation matrices in Supplementary Figure 3 for completeness sake, noting the strong negative correlations between cortical [^18^F]AV-1451 BP_ND_ and subcortical [^11^C]UCB-J BP_ND_. Third, we note that in PET studies of neurodegeneration with atrophy, grey matter volume loss can affect the interpretation of PET signals. However, synaptic loss in PSP and CBD occurs even in areas of the brain without discernible atrophy on MRI ^7,8^. Nonetheless, we used a stringent partial volume correction method (GTM) to minimise the effect of atrophy on our ligand cross-correlations. Of note, our data without partial volume correction yield similar results in all the main analyses (Supplementary Figure 1 and 2). Lastly, the cross-sectional design of this study limits the interpretation of the dynamic relationship between tau accumulation and synaptic loss. Although we include patients at various stages of their illness, a longitudinal design is necessary to test the dynamic relationship we propose, and the mediation of synaptic loss by progressive tauopathy.

In conclusion, we demonstrate a widespread positive association between [^18^F]AV-1451 and [^11^C]UCB-J binding in patients with symptomatic PSP and amyloid-negative corticobasal syndromes. Individual variability in this association correlates with disease severity. The complex relationship between tau accumulation and synaptic density *in vivo* may explain changes in cognitive and motor physiology. We hope that these insights will inform the design of new clinical trials to arrest PSP and CBD.

## Acknowledgements

The authors thank the research participants and caregivers, the staff at the Wolfson Brain Imaging Centre, and at the Cambridge Centre for Parkinson-Plus. We thank the NIHR Cambridge Biomedical Research Centre for support. We thank UCB Pharma, and Avid (Lilly) for providing the precursor for [^11^C]UCB-J and [^18^F]AV-1451 synthesis, respectively. NH and JBR had full access to all the data in the study and take responsibility for the integrity of the data and the accuracy of the data analysis.

## Abbreviations

PSP: Progressive Supranuclear Palsy – Richardson’s Syndrome
CBD: Corticobasal Degeneration
CBS: Corticobasal Syndrome
PSPRS: PSP Rating Scale

## References

1. Bigio EH, Vono MB, Satumtira S, et al. Cortical synapse loss in progressive supranuclear palsy. J Neuropathol Exp Neurol. 2001;60(5):403–410. doi:10.1093/jnen/60.5.403

2. Lipton AM, Munro Cullum C, Satumtira S, et al. Contribution of asymmetric synapse loss to lateralizing clinical deficits in frontotemporal dementias. Arch Neurol. 2001;58(8):1233–1239. doi:10.1001/archneur.58.8.1233

3. Clare R, King VG, Wirenfeldt M, Vinters H V. Synapse loss in dementias. J Neurosci Res. 2010;88(10):2083–2090. doi:10.1002/jnr.22392

4. DeKosky ST, Scheff SW. Synapse loss in frontal cortex biopsies in Alzheimer’s disease: Correlation with cognitive severity. Ann Neurol. 1990;27(5):457–464. doi:10.1002/ana.410270502

5. Terry RD, Masliah E, Salmon DP, et al. Physical basis of cognitive alterations in alzheimer’s disease: Synapse loss is the major correlate of cognitive impairment. Ann Neurol. 1991;30(4):572–580. doi:10.1002/ana.410300410

6. Jacobsen JS, Wu C-C, Redwine JM, et al. Early-Onset Behavioral and Synaptic Deficits in a Mouse Model of Alzheimer’s Disease. Vol 103.; 2006. www.pnas.orgcgidoi10.1073pnas.0600948103.

7. Holland N, Jones PS, Savulich G, et al. Synaptic Loss in Primary Tauopathies Revealed by [11C]UCB-J Positron Emission Tomography. Mov Disord. 2020;35(10):1834–1842. doi:10.1002/mds.28188

8. Mak E*, Holland, N* Jones, S, Savulich G, Low A, Malpetti M, Kaalund S, Passamonti L, Rittman T, Romero-Garcia R, Manavaki R, Williams G, Hong Y, Fryer T, Aigbirhio F, O’Brien J** RJ. In vivo coupling of dendritic complexity with presynaptic density in primary tauopathies. Neurobiol Aging. 2021:Accepted for publication.

9. DeVos SL, Corjuc BT, Oakley DH, et al. Synaptic tau seeding precedes tau pathology in human Alzheimer’s disease brain. Front Neurosci. 2018. doi:10.3389/fnins.2018.00267

10. Polanco JC, Li C, Durisic N, Sullivan R, Götz J. Exosomes taken up by neurons hijack the endosomal pathway to spread to interconnected neurons. Acta Neuropathol Commun. 2018;6(1):10. doi:10.1186/s40478-018-0514-4

11. Menkes-Caspi N, Yamin HG, Kellner V, Spires-Jones TL, Cohen D, Stern EA. Pathological tau disrupts ongoing network activity. Neuron. 2015;85(5):959–966. doi:10.1016/j.neuron.2015.01.025

12. Kaniyappan S, Chandupatla RR, Mandelkow EM, Mandelkow E. Extracellular low-n oligomers of tau cause selective synaptotoxicity without affecting cell viability. Alzheimer’s Dement. 2017;13(11):1270–1291. doi:10.1016/j.jalz.2017.04.002

13. Jiang S, Wen N, Li Z, et al. Integrative system biology analyses of CRISPR-edited iPSC-derived neurons and human brains reveal deficiencies of presynaptic signaling in FTLD and PSP. Transl Psychiatry. 2018;8(1):265. doi:10.1038/s41398-018-0319-z

14. Spires-Jones TL, Hyman BT. The Intersection of Amyloid Beta and Tau at Synapses in Alzheimer’s Disease. Neuron. 2014;82(4):756–771. doi:10.1016/j.neuron.2014.05.004

15. Kopeikina KJ, Polydoro M, Tai HC, et al. Synaptic alterations in the rTg4510 mouse model of tauopathy. J Comp Neurol. 2013;521(6):1334–1353. doi:10.1002/cne.23234

16. Mukaetova-Ladinska EB, Garcia-Siera F, Hurt J, et al. Staging of cytoskeletal and β-amyloid changes in human isocortex reveals biphasic synaptic protein response during progression of Alzheimer’s disease. Am J Pathol. 2000;157(2):623–636. doi:10.1016/S0002-9440(10)64573-7

17. Vanhaute H, Ceccarini J, Michiels L, et al. In vivo synaptic density loss is related to tau deposition in amnestic mild cognitive impairment. Neurology. June 2020:10.1212/WNL.0000000000009818. doi:10.1212/WNL.0000000000009818

18. Malpetti M, Kievit RA, Passamonti L, et al. Microglial activation and tau burden predict cognitive decline in Alzheimer’s disease. doi:10.1093/brain/awaa088

19. Höglinger GU, Respondek G, Stamelou M, et al. Clinical diagnosis of progressive supranuclear palsy: The movement disorder society criteria. Mov Disord. 2017;32(6):853–864. doi:10.1002/mds.26987

20. Alexander SK, Rittman T, Xuereb JH, Bak TH, Hodges JR, Rowe JB. Validation of the new consensus criteria for the diagnosis of corticobasal degeneration. J Neurol Neurosurg Psychiatry. 2014;85(8):923–927. doi:10.1136/jnnp-2013-307035

21. Kovacs GG, Lukic MJ, Irwin DJ, et al. Distribution patterns of tau pathology in progressive supranuclear palsy. Acta Neuropathol. May 2020:1–21. doi:10.1007/s00401-020-02158-2

22. Rösler TW, Tayaranian Marvian A, Brendel M, et al. Four-repeat tauopathies. Prog Neurobiol. 2019;180:101644. doi:10.1016/j.pneurobio.2019.101644

23. Zhou L, McInnes J, Wierda K, et al. Tau association with synaptic vesicles causes presynaptic dysfunction. Nat Commun. 2017;8(May):1–13. doi:10.1038/ncomms15295

24. Cope TE, Rittman T, Borchert RJ, et al. Tau burden and the functional connectome in Alzheimer’s disease and progressive supranuclear palsy. Brain. 2018;141(2):550–567. doi:10.1093/brain/awx347

25. Finnema SJ, Nabulsi NB, Eid T, et al. Imaging synaptic density in the living human brain. Sci Transl Med. 2016;8(348). doi:10.1126/scitranslmed.aaf6667

26. Bajjalieh SM, Peterson K, Linial M, Scheller RH. Brain Contains Two Forms of Synaptic Vesicle Protein 2. Vol 90.; 1993. doi:10.1073/pnas.90.6.2150

27. Burgos N, Cardoso MJ, Thielemans K, et al. Attenuation correction synthesis for hybrid PET-MR scanners: Application to brain studies. IEEE Trans Med Imaging. 2014. doi:10.1109/TMI.2014.2340135

28. Prados F, Cardoso MJ, Burgos N, et al. NiftyWeb: web based platform for image processing on the cloud. In: 24th Scientific Meeting and Exhibition of the International Society for Magnetic Resonance in Medicine (ISMRM). ; 2016.

29. Manavaki R, Hong Y, Fryer TD. Brain MRI coil attenuation map processing for the GE SIGNA PET/MR: Impact on PET image quantification and uniformity. IEEE Nucl Sci Symp Med Imaging Conf Proceedings. 2019.

30. Avants BB, Epstein CL, Grossman M, Gee JC. Symmetric diffeomorphic image registration with cross-correlation: Evaluating automated labeling of elderly and neurodegenerative brain. Med Image Anal. 2008;12(1):26–41. doi:10.1016/j.media.2007.06.004

31. Rousset OG, Ma Y, Evans AC. Correction for partial volume effects in PET: Principle and validation. J Nucl Med. 1998.

32. Wu Y, Carson RE. Noise reduction in the simplified reference tissue model for neuroreceptor functional imaging. J Cereb Blood Flow Metab. 2002. doi:10.1097/01.WCB.0000033967.83623.34

33. Koole M, van Aalst J, Devrome M, et al. Quantifying SV2A density and drug occupancy in the human brain using [11 C]UCB-J PET imaging and subcortical white matter as reference tissue. Eur J Nucl Med Mol Imaging. 2019;46(2):396–406. doi:10.1007/s00259-018-4119-8

34. Rossano S, Toyonaga T, Finnema SJ, et al. Assessment of a white matter reference region for 11C-UCB-J PET quantification. J Cereb Blood Flow Metab. 2019. doi:10.1177/0271678X19879230

35. Passamonti L, Rodríguez PV, Hong YT, et al. 18F-AV-1451 positron emission tomography in Alzheimer’s disease and progressive supranuclear palsy. Brain. 2017;140(3):781–791. doi:10.1093/brain/aww340

36. Klunk WE, Koeppe RA, Price JC, et al. The Centiloid project: Standardizing quantitative amyloid plaque estimation by PET. Alzheimer’s Dement. 2015;11(1):1-15.e4. doi:10.1016/j.jalz.2014.07.003

37. Jack CR, Wiste HJ, Weigand SD, et al. Defining imaging biomarker cut points for brain aging and Alzheimer’s disease. Alzheimer’s Dement. 2017;13(3):205–216. doi:10.1016/j.jalz.2016.08.005

38. Leuzy A, Chiotis K, Lemoine L, et al. Tau PET imaging in neurodegenerative tauopathies—still a challenge. Mol Psychiatry. 2019;24(8):1112–1134. doi:10.1038/s41380-018-0342-8

39. Dickson DW, Kouri N, Murray ME, Josephs KA. Neuropathology of frontotemporal lobar degeneration-Tau (FTLD-Tau). In: Journal of Molecular Neuroscience. Vol 45. NIH Public Access; 2011:384–389. doi:10.1007/s12031-011-9589-0

40. Jadhav S, Cubinkova V, Zimova I, et al. Tau-mediated synaptic damage in Alzheimer’s disease. Transl Neurosci. 2015;6(1):214–226. doi:10.1515/tnsci-2015-0023

41. Kneynsberg A, Combs B, Christensen K, Morfini G, Kanaan NM. Axonal degeneration in tauopathies: Disease relevance and underlying mechanisms. Front Neurosci. 2017;11(OCT):572. doi:10.3389/fnins.2017.00572

42. Vogels T, Murgoci A-N, Hromádka T. Intersection of pathological tau and microglia at the synapse. doi:10.1186/s40478-019-0754-y

43. Kovacs GG. Astroglia and Tau: New Perspectives. Front Aging Neurosci. 2020;12:96. doi:10.3389/fnagi.2020.00096

44. Clavaguera F, Bolmont T, Crowther RA, et al. Transmission and spreading of tauopathy in transgenic mouse brain. Nat Cell Biol. 2009;11(7):909–913. doi:10.1038/ncb1901

45. Clavaguera F, Akatsu H, Fraser G, et al. Brain homogenates from human tauopathies induce tau inclusions in mouse brain. Proc Natl Acad Sci U S A. 2013;110(23):9535–9540. doi:10.1073/pnas.1301175110

46. Malpetti M, Passamonti L, Jones PS, et al. Neuroinflammation predicts disease progression in progressive supranuclear palsy. medRxiv. May 2020:http://dx.doi.org/10.1136/jnnp-2020-325549. doi:http://dx.doi.org/10.1136/jnnp-2020-325549

47. Palleis C, Sauerbeck J, Beyer L, et al. In Vivo Assessment of Neuroinflammation in<scp>4-Repeat</scp> Tauopathies. Mov Disord. November 2020:mds.28395. doi:10.1002/mds.28395

48. Malpetti M, Passamonti L, Rittman T, et al. Neuroinflammation and tau co[localize in vivo in progressive supranuclear palsy. Ann Neurol. September 2020:ana.25911. doi:10.1002/ana.25911

49. Gerhard A, Trender-Gerhard I, Turkheimer F, Quinn NP, Bhatia KP, Brooks DJ. In vivo imaging of microglial activation with [11C]-PK11195 PET progresive supranuclear palsy. Mov Disord. 2006;21(1):89–93. doi:10.1002/mds.20668

50. Gerhard A, Watts J, Trender-Gerhard I, et al. In vivo imaging of microglial activation with [11C](R -PK11195 PET in corticobasal degeneration. Mov Disord. 2004;19(10):1221–1226. doi:10.1002/mds.20162

51. Metaxas A, Thygesen C, Briting SRR, Landau AM, Darvesh S, Finsen B. Increased Inflammation and Unchanged Density of Synaptic Vesicle Glycoprotein 2A (SV2A) in the Postmortem Frontal Cortex of Alzheimer’s Disease Patients. Front Cell Neurosci. 2019;13. doi:10.3389/fncel.2019.00538

52. Padovani A, Borroni B, Brambati SM, et al. Diffusion tensor imaging and voxel based morphometry study in early progressive supranuclear palsy. J Neurol Neurosurg Psychiatry. 2006;77(4):457–463. doi:10.1136/jnnp.2005.075713

53. Borroni B, Garibotto V, Agosti C, et al. White matter changes in corticobasal degeneration syndrome and correlation with limb apraxia. Arch Neurol. 2008;65(6):796–801. doi:10.1001/archneur.65.6.796

