## Supplementary figures 1-3 for "Molecular pathology and synaptic loss in primary tauopathies: [^18^F]AV-1451 and [^11^C]UCB-J PET study"

Supplementary Figures = 3

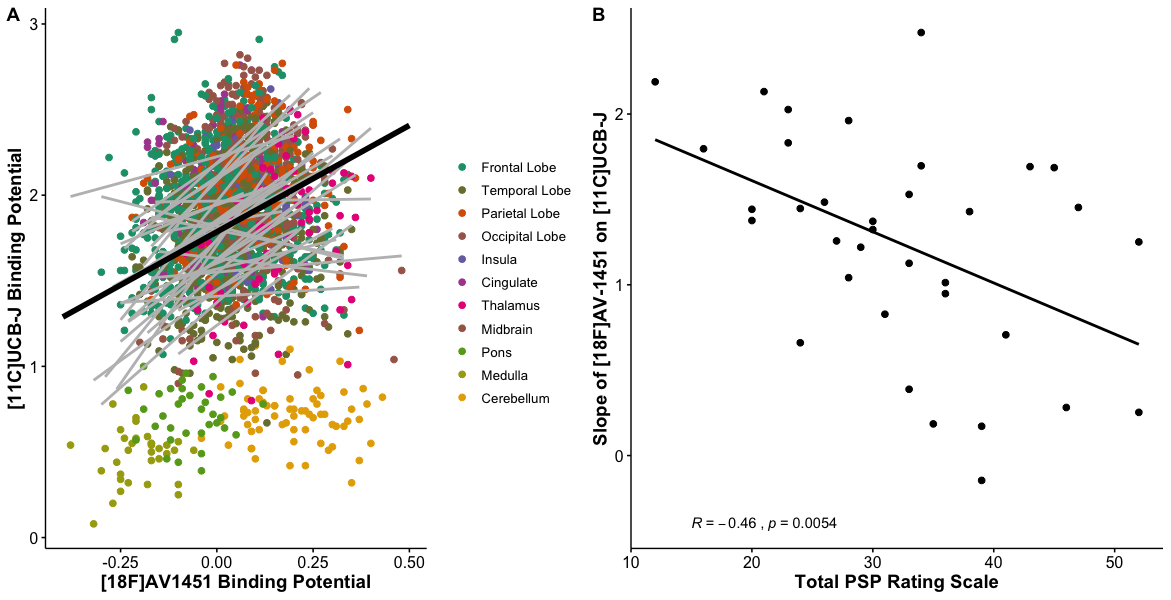

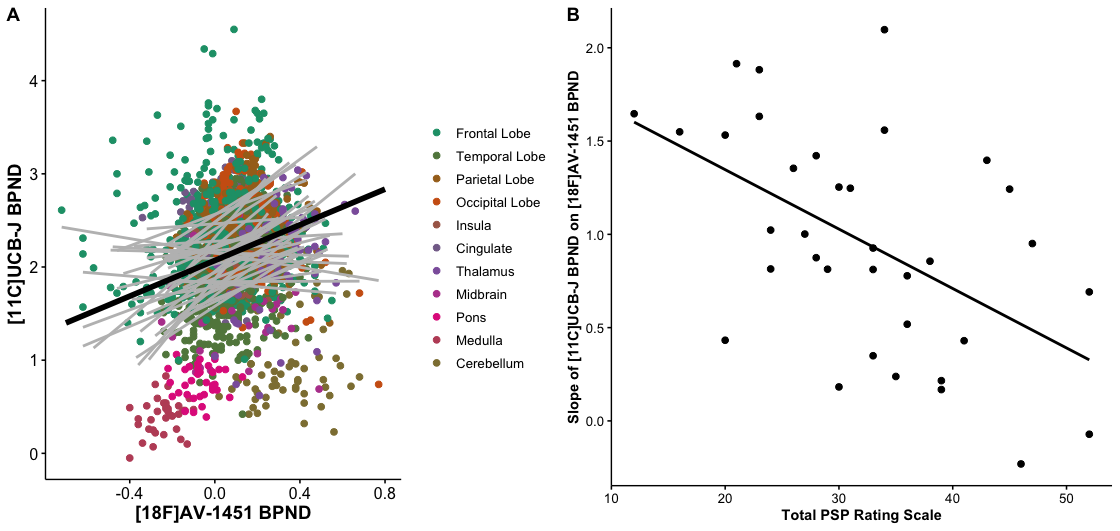

**Supplementary Figure 1.** **The association between synaptic density ([^11^C]UCB-J) and molecular pathology ([^18^F]AV-1451) is a function of disease severity.** A) Scatter plot [^11^C]UCB-J and [^18^F]AV-145 **partial volume uncorrected** BP_ND_ from 23 patients with PSP and 12 patients with amyloid-negative corticobasal syndrome . Each grey line in A represents a patient’s data across 73 regions of interest (excluding those with previously reported off-target binding, i.e. basal ganglia and substantia nigra); the black line illustrates the overall model fit from equation 2. B) The slope for each individual (i.e. each grey line in A) is negatively correlated with disease severity (as measured with the PSP rating scale); R= -0.6, p<0.005.

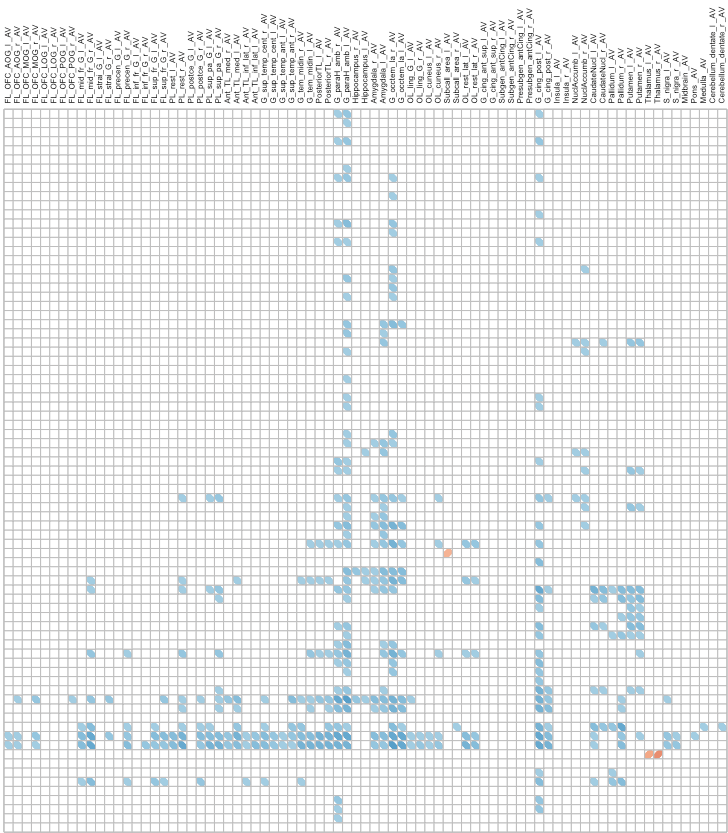

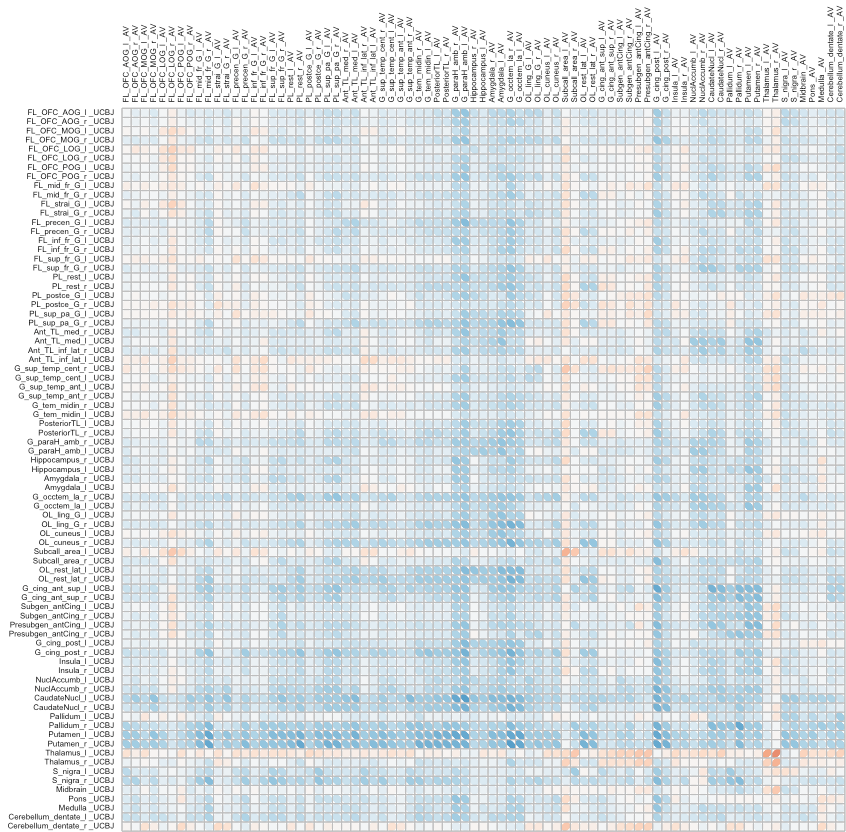

Frontal

Parietal

Temporal

Occipital

BG

BS

Cingulate

Frontal

Parietal

Temporal

Occipital

BG

BS

Cingulate

Frontal

Parietal

Cingulate

Temporal

Occipital

BS

BG

Frontal

Parietal

Temporal

Occipital

BG

BS

Cingulate

Frontal

Parietal

Temporal

Occipital

BG

BS

Cingulate

Frontal

Parietal

Cingulate

Temporal

Occipital

BS

BG

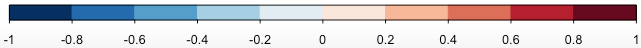

[^18^F]AV-1451

[^18^F]AV-1451

[^11^C]UCB-J

**Supplementary figure 2.** **Cortical molecular pathology is negatively correlated with subcortical synaptic density.** Correlation between [^18^F]AV-1451 BP_ND_ in a source region (horizontal axis) and [^11^C]UCB-J BP_ND_ in a target region (vertical axis), across 79 regions of interest in patients, using **partial volume uncorrected binding potentials**. The black box in the lower left quadrant focuses on cortical [^18^F]AV-1451 BP_ND_ and subcortical [^11^C]UCB-J BP_ND_. **Significant correlations (at p<0.05, uncorrected for multiple comparisons) are outlined in the matrix on the right.**

Abbreviation: l: left, r:right, FL: Frontal Lobe, OFC: Orbitofrontal Cortex, AOG: Anterior Orbital Gyrus, MOG: Middle Orbital Gyrus, LOG: Lateral Orbital Gyrus, POG: Posterior Orbital Gyrus, mid_fr_G: Middle Frontal Gyrus, strai_G: Straight Gyrus, precen_G: Precentral Gyrus, inf_fr_G: Inferior Frontal Gyrus, sup_fr_G: Superior Frontal Gyrus; PL: Parietal Lobe, postce_G: Postcentral Gyrus, sup_pa_G: Superior Parietal Gyrus: TL: Temporal Lobe, Ant_med: Anterior Medial, Ant_inf_lat: Anterior Inferiolateral, "G_sup_temp_cent: Superior Temporal Gyrus – superior part, G_sup_temp_ant: Superior Temporal Gyrus- Anterior part, G_tem_midin_r: Middle and Inferior Temporal Gyrus, G_paraH_amb: Parahippocampal and ambient gyri, G_occtem_la: Occipitotemporal Gyrus – Lateral Part (Fusiform Gyrus), OL_ling_G_l: Lingual Gyrus, Subcall_area: Subcallosal Area, OL_rest_lat: Latera Remainder of Occipital Lobe, G_cing_ant_sup: Cingulate Gyrus Anterior Part, Subgen_antCing: Subgenual Frontal Lobe, Presubgen_antCing: Presubgenual Frontal Lobe, G_cing_post: Cingulate Gyrus – posterior part; BG: Basal Ganglia; BS: Brainstem

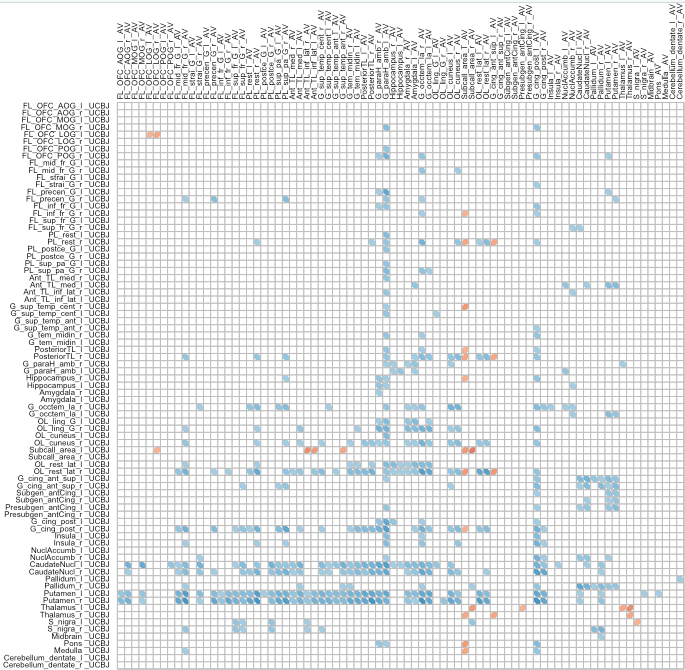

Frontal

Parietal

Temporal

Occipital

BG

BS

Cingulate

Frontal

Parietal

Temporal

Occipital

BG

BS

Cingulate

Frontal

Parietal

Cingulate

Temporal

Occipital

BS

BG

Frontal

Parietal

Temporal

Occipital

BG

BS

Cingulate

Frontal

Parietal

Temporal

Occipital

BG

BS

Cingulate

Frontal

Parietal

Cingulate

Temporal

Occipital

BS

BG

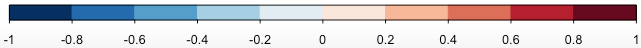

[^18^F]AV-1451

[^18^F]AV-1451

[^11^C]UCB-J

**Supplementary figure 3.** **Cortical molecular pathology is negatively correlated with subcortical synaptic density.** Correlation between [^18^F]AV-1451 BP_ND_ in a source region (horizontal axis) and [^11^C]UCB-J BP_ND_ in a target region (vertical axis), across 79 regions of interest in patients, using **GTM partial volume corrected binding potentials**. The black box in the lower left quadrant focuses on cortical [^18^F]AV-1451 BP_ND_ and subcortical [^11^C]UCB-J BP_ND_. **Significant correlations (at p<0.05, uncorrected for multiple comparisons) are outlined in the matrix on the right.**

Abbreviation: l: left, r:right, FL: Frontal Lobe, OFC: Orbitofrontal Cortex, AOG: Anterior Orbital Gyrus, MOG: Middle Orbital Gyrus, LOG: Lateral Orbital Gyrus, POG: Posterior Orbital Gyrus, mid_fr_G: Middle Frontal Gyrus, strai_G: Straight Gyrus, precen_G: Precentral Gyrus, inf_fr_G: Inferior Frontal Gyrus, sup_fr_G: Superior Frontal Gyrus; PL: Parietal Lobe, postce_G: Postcentral Gyrus, sup_pa_G: Superior Parietal Gyrus: TL: Temporal Lobe, Ant_med: Anterior Medial, Ant_inf_lat: Anterior Inferiolateral, "G_sup_temp_cent: Superior Temporal Gyrus – superior part, G_sup_temp_ant: Superior Temporal Gyrus- Anterior part, G_tem_midin_r: Middle and Inferior Temporal Gyrus, G_paraH_amb: Parahippocampal and ambient gyri, G_occtem_la: Occipitotemporal Gyrus – Lateral Part (Fusiform Gyrus), OL_ling_G_l: Lingual Gyrus, Subcall_area: Subcallosal Area, OL_rest_lat: Latera Remainder of Occipital Lobe, G_cing_ant_sup: Cingulate Gyrus Anterior Part, Subgen_antCing: Subgenual Frontal Lobe, Presubgen_antCing: Presubgenual Frontal Lobe, G_cing_post: Cingulate Gyrus – posterior part; BG: Basal Ganglia; BS: Brainstem
